# Assessing the impact of multiple comorbidities on fatal outcome in young COVID-19

**DOI:** 10.1101/2021.03.29.21254599

**Authors:** Paulino Monroy-Castillero, Eitan Friedman, Arturo Revuelta-Herrera, Arik Yochelis

## Abstract

A Bayesian analysis with the use of a rank-biserial correlation algorithm was applied to identify the impact of multiple comorbid conditions on fatal COVID-19 outcome in young adult cases (40-50 years). The demonstration was conducted for a publicly available database provided by the Mexican authority, in the absence of other alternative free-access repositories with information per patient. The methodology here proposed showed that even in the face of small sample sizes, it is possible to highlight deleterious synergistic comorbid conditions.

Young adult cases with COVID-19 and co-existing diabetes, obesity, hypertension, CRF, or COPD were found more likely to have a fatal outcome compared with having no co-morbidities (X2-6 times). With the methodology proposed, we show that having diabetes or hypertension in addition to CRF increased risk for mortality more than what is expected from independent effect (adverse synergistic effect), whereas in patients with obesity, the additional presence of diabetes or hypertension do not increase markedly the death risk due to COVID-19. Quantitative analysis of having two comorbidities highlights the combinations of morbid conditions that are more likely to be associated with fatal outcomes in younger adults COVID-19 cases in a clinically applicable manner.

The clinical implication of this method needs to be prospectively assessed.

## Introduction

The novel SARS-CoV-2 virus first described in China at the end of 2019 is the causative agent of COVID-19 pandemic.^1^ Up to December 13, 2020, there were 72,653,057 confirmed COVID-19 cases and 1,618,890 COVID-19 related deaths globally.^2^ While the majority of COVID-19 cases exhibit mild symptoms with a self-limiting clinical course and do not require hospitalization, a subset of cases develop severe disease hallmarked by bilateral pneumonia and acute respiratory distress requiring ventilation, with a minority of COVID-19 cases dying of the disease.^3,4^ By early estimates infection fatality ratio are ∼0.5%.^6^ Case fatality rate is hard to assess in real time as multiple confounders limit the ability to compare inter country rates (e.g., testing indications, population genetic makeup, associating a direct causal effect on COVID-19 as a cause of death and timing, associated comorbidities, population median age differences, and gender specific fatalities) to name a few. While age (above 65 years) and male gender adversely affect disease course and are associated with higher mortality rates, other comorbid conditions also play a role. Notably, hypertension, diabetes mellitus, chronic obstructive pulmonary disease (COPD), cardiovascular diseases, obesity, and chronic renal failure (CRF) are all comorbidities that are well established as associated with more adverse COVID-19 outcomes^7–9^ Specifically in Mexico by November 13, 2020, of 323,671 confirmed cases at the Mexican Institute of Social Security, hypertension (reported in 40.4% of cases), diabetes (33.1%) and obesity (21.7%) were more frequently noted in cases with fatal outcome.^10^ Identification and quantitation of risk factors associated with disease severity and/or mortality among COVID-19 patients is urgently needed, ideally using data analysis scheme that is based on actual data without any model driven assumptions.

COVID-19 patients who are younger than 60 years of age (and hence considered at low risk for developing severe disease and fatal outcome). ^11^ Thus, the ability to evaluate the contribution of risk factors associated with severe outcome in this age group is essential for early intervention in order to potentially improve prognosis and for priorization schemes for vaccine distribution. Most risk factor analyses studies have focused on a single factor and its effect on disease severity or mortality on all age groups, employing assumption-driven models to compensate for and extrapolate over missing data, such as a logistic regression-type approaches,^12–14^ while some others are based on artificial intelligence algorithms and provide a binary outcome (mortality or recovery). ^15^ Based on correlation with mortality rates, a common theme has emerged that diabetes mellitus, hypertension, and obesity are the most significant comorbidities associated with COVID 19 mortality.^7,10^ However, assessment of an individualized adverse disease course should not rely on retrospective analyses but rather assess these risks as a function of comorbidities in a prospective manner, data that is often incompletely collected and curated.^16^ Thus, a key component in identifying the relative contribution of each factor to adverse disease outcome and the possible interaction between factors affecting morality should be hypothesis-free methodology. Applying a generic Bayesian inference to COVID-19 Mexican real-time data cases, we show that it is possible to identify synergistic effects of comorbid conditions and compare their effects on mortality risk, even in the presence of incomplete data.

## Methods

### Data Source and Collection

Deidentified, open access, repository of data collected on all individuals who tested positive for SARS-CoV-2 by reverse transcription polymerase chain reaction (RT PCR) using one of several commercially available kits that were validated and approved for use by the Institute of Epidemiological Diagnosis and Reference (InDRE)^17^ in Mexico, were obtained from the web site of the Secretaría de Salud, Mexican Federal Government,^18^ (accessed December 10, 2020). This site contains all Mexican subjects tested for SARS CoV-2 in public institutions since February 2020. For each individual, the following data were obtained: age, gender, alive/deceased status, date of symptom onset, and any of the following, self-reported comorbidities: diabetes mellitus, chronic obstructive pulmonary disease (COPD), asthma, immunosuppression, hypertension (HTN), cardiovascular disease (CVD), obesity (BMI>30), chronic renal failure (CRF), and smoking status (never/ever). To avoid bias that may have been introduced by reporting delays of disease outcome (deaths or recovery), we only included individuals with symptom onset date earlier than November 11, 2020.

## Statistical Analysis

Given the incompleteness of data and lack of established knowledge on specific a priory probability, Bayesian methodology was chosen for inference.^19–23^ Here, we focus on factors that putatively affect the case fatality rate (CFR) by following the path outlined in Fig. 1: (i) using clinical data to substantiate the posterior probability distribution of the CFR, referred to as case fatality rate distribution (CFRD), and their respective relative risks (RR), and (ii) identifying synergistic effects in the presence of two comorbidities in a qualitative and a quantitative way, the latter by using the rank-biserial correlation (RBC). ^24^

**Figure 1:**
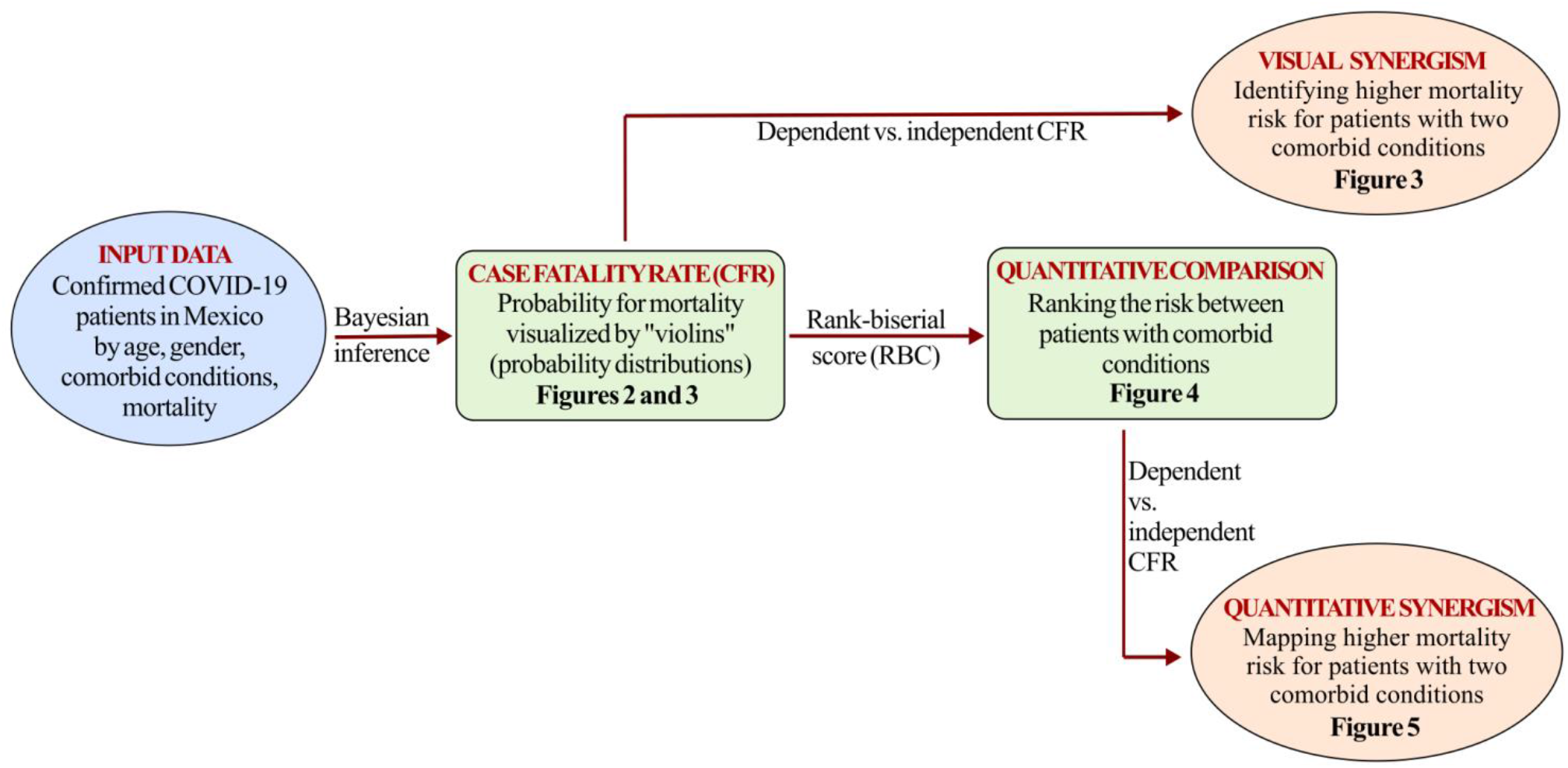
Flowchart of the conducted study.

## Case Fatality Rate (CFR) and Relative Risk (RR)

The complied data encompassed the total number of patients (*n*) and the number of deaths (*d*) by gender, age, and comorbid conditions. Using Bayesian inference^21^ data-based case fatality rate distribution (CFRD) which outlines the likelihood of the CFR was calculated. From a statistical view point, CFR is the true value, and CFRD is the so-called posterior probability distribution that displays less spreading along the vertical axis as *n* increases.^21–23^

Initially, CFR from COVID-19 cases by age group (at 5-year intervals) for individuals with no comorbid conditions were calculated (Fig. 2). While the “true” CFR remains unknown, the so-called “violins” in Fig. 2 outline the likelihood and precision of CFR. Next, the RR in a presence of a single comorbid condition for males and females separately, was calculated (Fig. 3). The likelihood of RR is again outlined by a distribution of the CFRD of patients that is normalized by the CFRD of cases with no comorbid condition, as is shown in Fig. 2. Fig. 3 outlines the CFRD of patients having two comorbid conditions. In order to identify any possible deleterious comorbidity synergism, the combined “theoretical” CFRD was plotted as if both comorbidities have no synergistic effect but rather an additive effect (gray color violins in Fig. 3).

**Figure 2:**
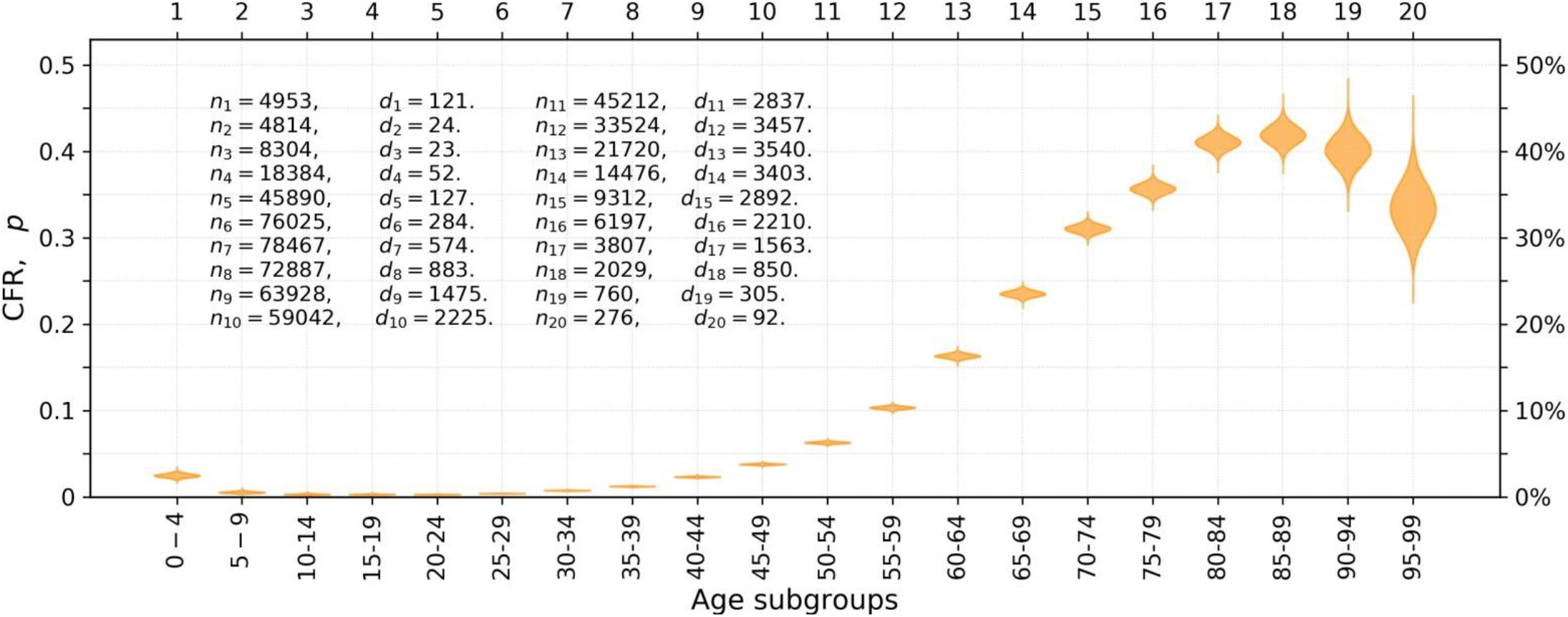
Age dependent risk for developing adverse disease course in the absence of comorbid conditions. Case fatality rate (CFR) likelihood outlined by “violins” (posterior beta distributions) of individuals within age intervals. The violins are constructed using a standard Bayesian update procedure^22,23^ by starting from a uniform prior distribution and by incorporating data about the number of patients, *n*_*i*_, and the number of deaths, *d*_*i*_, in each subgroup (*i* = 1, …, 20). Specifically, we use a large sample of 50,000 random variates (realizations via Monte Carlo method^25,26^) from the beta distribution, *B*(*p*; *α, β*), where *α* = *d*_*i*_ + 1 and *β* = *n*_*i*_ − *d*_*i*_ + 1. Violin’s elongation along the vertical axis indicates the precision in the probability of CFR, i.e., the smaller vertical extent is the closer is the result to “true” value.

**Figure 3:**
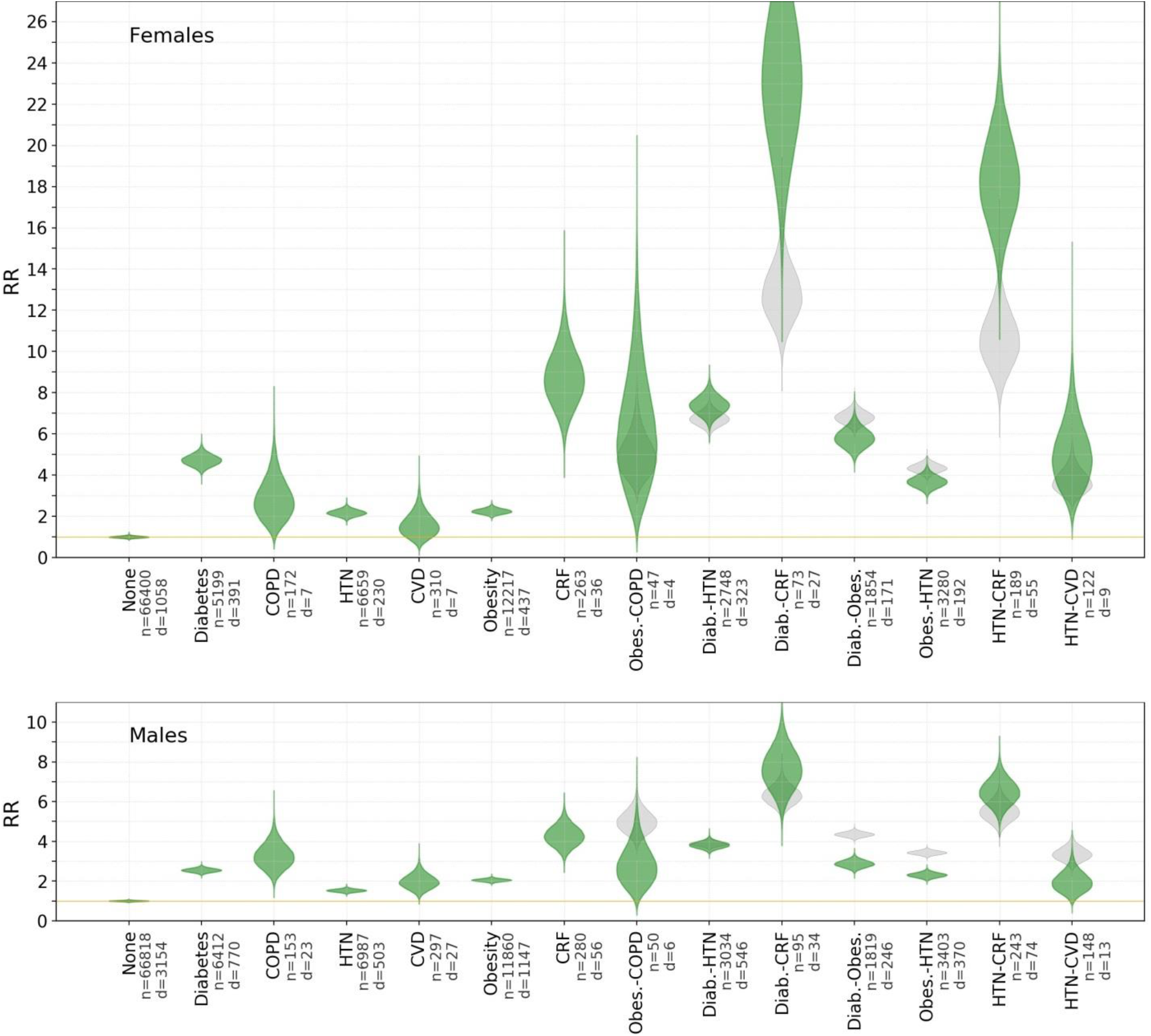
Relative risks (RR) for comorbid conditions. Green color violins depict the RR distributions obtained from the data while the gray violins depict a reference of distributions for conditions of two independent comorbid conditions having additive contributions. Specifically, computation of the relative risk distribution (RRD) of each subgroup is obtained by Bayesian update^23^ (as in Fig. 2) but for RR = *p*/*p*_*ref*_, where *p* and *p*_*ref*_ are random variates taken from the data for comorbid condition forming *B*_*data*_(*p*) and a CFRD for the reference subgroup in the absence of comorbid condition *B*_*ref*_(*p*). For a pair of independent comorbidities, gray shaded violins, we define the distribution with additive property, CFRD_add_, that is generated via the Monte Carlo method that randomly samples the probability for mortality *p*_add_ = 1 − (1 − *p*_comorbid 1_)(1 − *p*_comorbid 2_), where *p*_comorbid 1_ and *p*_comorbid 2_ are random variates of each comorbid condition CFRD separately.^26^ The results represent data for males and females at age interval 40-50 years.

## Rank-biserial correlation (RBC)

While it is possible to visualize the synergistic effects in patients with two comorbid conditions (Fig. 3), the calculated CFRD can also be used to get a quantitative comparative ranking. To that end, the RBC can be used to compare the risk of two patients with distinct comorbid conditions. Formally, the RBC is a correlation effect size for the Mann-Whitney U test. ^24^ Then, we set a threshold |RBC| ≥ 0.9 which provides the region for which it is possible to identify with a significance of 10%, who is at higher risk. Several examples are shown in Fig. 4. Finally, we also use RBC to detect some synergetic effect of two comorbidities, as shown in Fig. 5.

**Figure 4:**
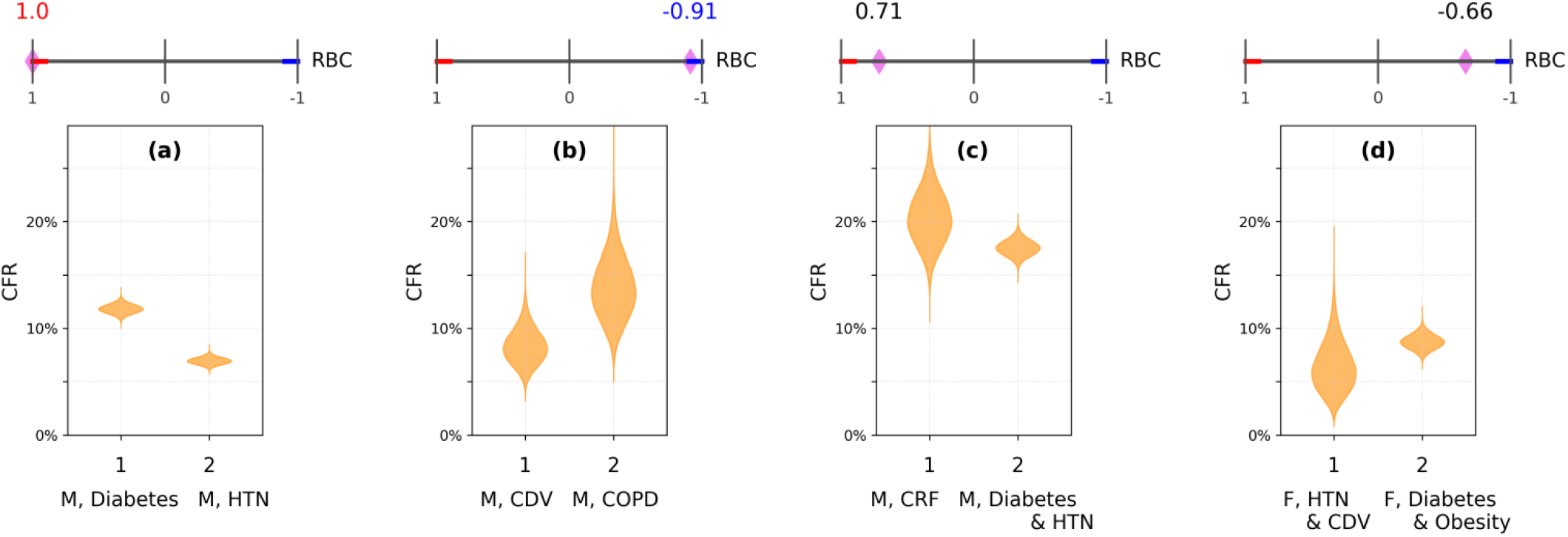
Risk ranking of patients with comorbid conditions. Selected cases of patients (labeled as 1 and 2) with distinct comorbid conditions, shown by the CFRD. The results taken for males (M) and females (F) at age interval 40-50 years. The top bar indicates the rank-biserial correlation (RBC) performed from a Monte Carlo sampling of each CFRD, where the supra-thresholds RBC ≥ 0.9 and RBC ≤ −0.9 are marked by red and blue colors, respectively. ^24^ The diamond symbol marks the RBC value according to: RBC(*B*_1_, *B*_2_) = *P*(*p*_1_ > *p*_2_) − *P*(*p*_2_ > *p*_1_), where *P*(*p*_1_ > *p*_2_) is the probability that the CFR for patient 1 (*p*_1_) is higher than CFR for patient 2 (*p*_2_) based on their respective CFRD, which are represented by the yellow violins. *P*(*p*_2_ > *p*_1_) = 1 − *P*(*p*_1_ > *p*_2_).

**Figure 5:**
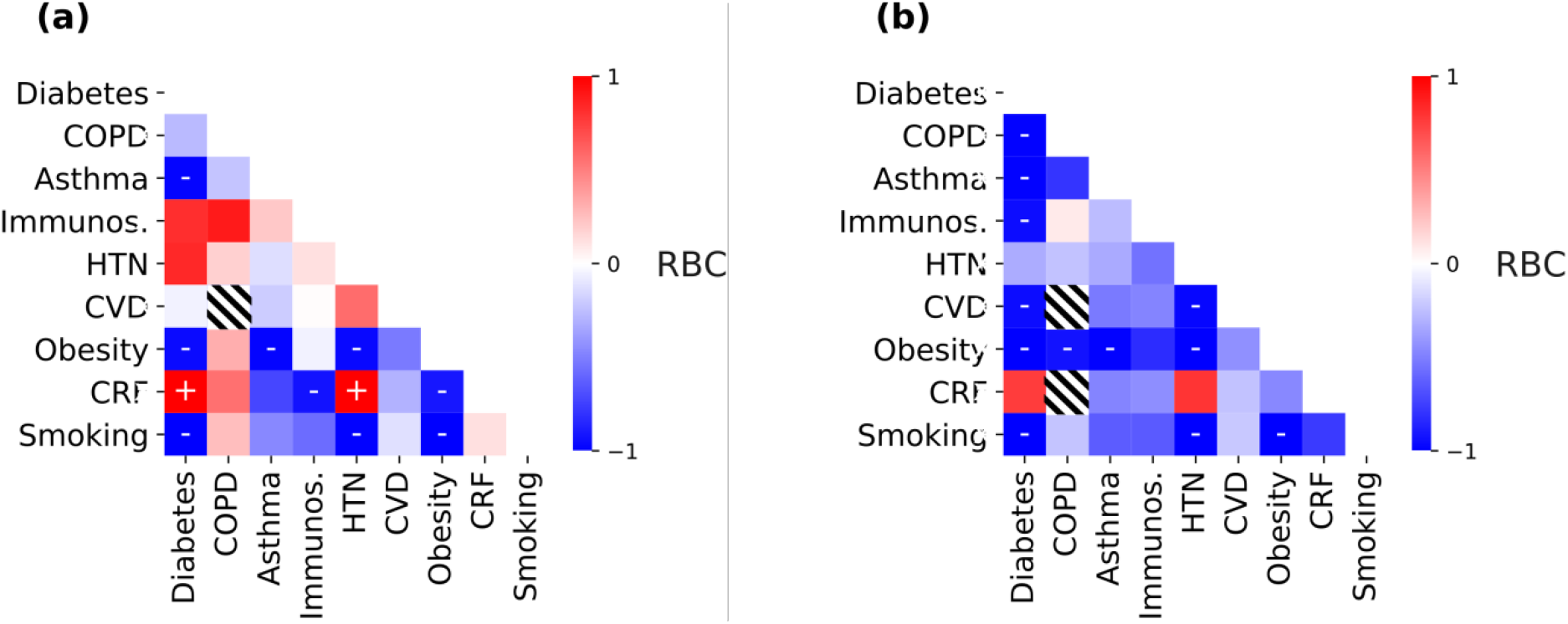
Identification of synergism between two comorbid conditions. Mapping of RBC(*B*_1_, *B*_2_), where *B*_1_ is related to a pair of concurrent comorbid conditions and *B*_2_ = *B*_*add*_, is the additive CFRD of the respective pair, see also Fig. 4. The RBC and the respective supra-thresholds values (together with ‘+’ and ‘-’, respectively), are computed and marked as in Fig. 4; we do not reflect and in fact ignore the comparisons that yield sub-threshold score. Half-filled squares indicate the absence of sufficient data (i.e., subgroups with less than 3 patients). (a) Females. (b) Males. Both at age interval 40-50 years.

## Results

### Cohort Description

By December 10, 2020, 1,174,784 individuals (578,791 females and 595,993 males) who tested positive for SARS-CoV-2 in Mexico were available for analysis from the Secretaría de Salud, Mexican Federal Government database.^18^ Eligible individuals (with symptom onset date earlier than November 11, 2020) were subdivided into three groups: 570,094 individuals with no known or reported comorbidities (as enumerated in the methods section), 264,703 individuals with a single morbid condition, and 121,774 individuals with two comorbidities. Noteworthy, the current analysis included only cases with up to two comorbidities whereas individuals with more than 2 comorbid conditions were excluded. Given the major effect of age on mortality and the paucity of data on the effect of comorbidities on mortality in younger cases, we focused on the seemingly less susceptible age group of 40 to 50 years, that encompassed 224,616 patients (11,219 deaths) in total. For that subgroup, of 110,561 females 11,219 deaths were reported, and 7,866 of 114,055 males died.

### Relative risk by comorbidity status

#### Effect of Single Comorbid Condition on Relative Risk (RR)

As shown in Fig. 3, diabetes mellitus, CRF, and obesity were associated with a mean mortality RR of X2-3 in males, and higher means in females. Furthermore, while the RR distributions (RRD) for diabetes mellitus and obesity showed restricted distributions, the RRD for CRF and COPD exhibit smeared RRD (i.e., elongated along the y-axis violins).

This implies that there is more uncertainty as to the RR conferred by the CRF compared with diabetes and obesity. In addition, COPD and hypertension were also associated with an increased RR in females that is more pronounced compared with the effects of these factors in males. Overall, RRD in females are more elongated (hence more uncertain) and have a mean of higher values compared with males.

#### Effect of Two Comorbid Conditions on RR

As shown in Fig. 3, having more than one comorbid condition in both genders may affect the likelihood of having higher RR. For example, having both CRF and diabetes mellitus increases the RR and the RRD spreads, implying an adverse effect, along with increased uncertainty due to the limited number of cases displaying these two conditions. Hypothetically, RRD in cases reporting two comorbidities could have independent additive effects, indicated by the gray color “violins” in Fig. 3. If the actual RRD (green violins in Fig. 3) are above those outlined by the grey violins, with no or minor overlap then it may indicate a synergistic deleterious impact on disease outcome. Conversely, comorbid conditions may have a seemingly “protective effect” on mortality in COVID-19 cases, namely the RRD is lower than the independent combination. Specific examples of both adverse and seemingly protective effects are shown in Fig. 3.

While the impact of having two comorbidities can be visually deduced from Fig. 3, the analysis may also enable quantitative differences, as shown by pair-wise combination in Fig. 5. Here, the information corresponds only to a dichotomous depiction of the possibility of synergy while disregarding relative risk values (see Methods section for details). Specifically, the results show that only for a few combinations of comorbidities it is possible to determine a clear-cut synergistic effect (e.g., diabetes or HTN with CRF), information that quantifies the visual comparison in Fig. 3. Notably, the combinations of obesity with either asthma, diabetes or HTN, show a non-synergistic and even a seemingly protective effect.

## Discussion

The analysis used herein is assumption-free and relies only on statistical inference of the available data. Specifically, it enables to quantitatively compare between two risk profiles (Fig. 4), a seemingly useful aid in clinical decision making. Although the statistical sampling used herein is still not big enough to establish conclusive determination whether some of the comorbidities pairs are associated or not with synergistic deleterious effects, is indeed enough to assert that there are combinations that deserve special attention: CRF with diabetes mellitus or HTN significantly increase RR and RRD for mortality. Conversely, the combinations of obesity with hypertension or diabetes mellitus seem not to have a synergistic effect on mortality RR. On the other hand, diabetes with HTN seems to present an independent combination effect; although is not synergistic according to our definition, the increase of risk is quite substantial to be highlighted for medical purposes. The mechanisms that underlie the observed synergistic or lack of such effect of combined comorbidities, remain unknown. A major limitation of this study that can be easily improved by worldwide information sharing is the incompleteness of data collection. Another limitation may be inherent to the fact that the data were collected retrospectively and pertain to a single country with distinct genetic makeup. Lastly, there are a limited number of cases in some categories (e.g., COPD females n=172, d=7, and males n=153, d=23 or CRF females n=263, d=36, and males n=280, d=56).

Unlike other available predictive risk methods that rely on estimations of parametric models from the whole dataset to predict risk profiles,^12–14^ the method applied and described herein used solely data of subgroups that are of interest, without further assumptions, and provides an order relation based on statistical comparison of risks. In the meta-analysis of observational studies by Parohan and coworkers^3^ risk factors (e.g., hypertension, diabetes mellitus, obesity, cerebrovascular stroke (CVS), COPD) individually were associated with an odds ratio (OR) of 1.5-4.59 for fatal COVID-19 outcome. Additional analyses refined these risks by individual risk factors (some of which are shown in Supplementary Table 1). Specifically in Mexico, a study encompassing 51,633 subjects with SARS-CoV-2 and 5,332 deaths, fatal outcome risk factors include early-onset diabetes mellitus, obesity, COPD, advanced age, HTN, immunosuppression, and CRF.^14^ Yet, it had been difficult to clinically apply the known risk factors in order to prioritize care for COVID-19 patients, due to the inaccuracy of assessing the relative contribution of each risk factor and their possible interactions as determinants of deleterious disease outcome.

This methodology can be employed clinically with a real-time updating to stratify cases of different mortality risks in the emergence of epidemics and to guide allocation of medical resources or prioritizing vaccine distribution, if the ability of this tool can be ascertained and validated prospectively.

The accuracy of the tool developed reported herein should be validated in other countries and using more data with more cases in each of the morbid conditions categories. In addition, the accuracy of this tool should be tested in a prospective study design and its added value as a clinical support decision tool should be evaluated.

## Data Availability

The authors attest that they are willing to share the data presented herein upon request.

