## Supplementary material for "Assessing the impact of multiple comorbidities on fatal outcome in young COVID-19"

**Supplementary Table 1 – Effect of comorbid conditions on fatality in COVID-19 cases.** Here RR is relative risk, OR is odds ratio, CI is confidence interval, and CVA is cerebrovascular accident. Other abbreviations are defined in the text.

| Risk factor | # cases | # fatalities | OR/RR for fatal outcome | 95% CI | ref # |
|---|---|---|---|---|---|
| **Hypertension** | **2389** | **151** | **3.48 (all) (OR)** | **1.72-7.08** | |
| | | | **6.43 (<50 years)** | **3.4-12.17** | |
| | | | **2.66 (>50 years)** | **1.27-5.57** | **1** |
| | **6560** | **799** | **2.21 (RR)** | **1.74-2.81** | **2** |
| **Diabetes Mellitus** | **1985** | **423** | **2.12 (RR)** | **1.44-3.11** | 3 |
| **CVS** | **1170** | **75** | **2.25 (RR)** | **1.53-3.29** | |
| **CVA** | **1337** | **75** | **2.25 (RR)** | **1.51-3.36** | **4** |
| **COPD/ non COPD** | **10/157** | **6/86** | **1.1 (RR)** | **0.6-1.8** | **5** |
